# Altered resting state EEG microstate dynamics in acute-phase pediatric mild traumatic brain injury

**DOI:** 10.1101/2024.10.26.24316185

**Authors:** Sahar Sattari, Samir Damji, Julianne McLeod, Maryam S. Mirian, Lyndia C. Wu, Naznin Virji-Babul

## Abstract

**Objective:** Sport-related concussion presents significant diagnostic and monitoring challenges, especially in youth populations. This study investigates the potential of EEG microstate analysis as a tool for assessing acute-phase brain activity changes in adolescent male athletes following a concussion. We analyzed resting-state EEG data from 32 participants in a between-subjects design, comparing participants with acute concussion (within two weeks of injury) to an age- and sex-matched sample with no reported history of concussion.

**Methodology:** We applied a modified k-means clustering algorithm to group resting-state EEG topographical maps into seven clusters, with each cluster represented by one of the canonical microstate classes (A-G). Average duration, occurrence rate, and time coverage for each microstate were extracted.

**Results:** Statistically significant differences in mean duration, occurrence rate, and time coverage of microstates B and E were observed. Specifically, the mean duration, occurrence and time coverage of microstate E showed a significant decrease in the concussed cohort in comparision to the controls (*p* < 0.001). In addition, the mean duration, occurrence rate and time coverage was higher in the concussed cohort in comparision with the healthy cohort (*p* = 0.003). A significant negative linear relationship was found between microstate E and symptom severity (*p = 0.006, F = 15.72*).

**Discussion:** These results suggest that mild traumatic brain injury may disrupt the dynamic interaction of large-scale brain networks, hinting at potential biomarkers of injury. This study may help to inform future work on objective, brain-based tools for diagnosis and recovery assessment in concussed adolescents. Further research in larger, more diverse populations is necessary to validate these potential biomarkers.

## Introduction

The neurophysiological and neurobiological changes due to mild traumatic brain injury (mTBI; hereafter used synonymously with concussion) are not well understood. Concussion results from biomechanical forces exerted to the skull and the subsequent shearing and stretching of brain tissue; although this injury is particularly prevalent, concussion diagnosis remains a challenge due to its largely subjective nature and the absence of definitive clinical tests [1]. There is a need for objective, brain-based diagnostic methods that can reliably indicate the occurrence of a concussion [2,3]. The dynamic and complex nature of concussion injury is of particular concern in adolescents due to the complexity and heterogeneity of neurodevelopment.

In Canada, concussion incidence is highest among adolescents aged 12 to 19 compared to other age brackets across the lifespan; this result is corroborated by a similar epidemiological investigation of the US population [4,5]. It should be noted that most concussion injuries in this age group are sustained during sport (hockey, rugby, and football in particular) and other physical activities [4].

Adolescents are particularly vulnerable to the neuropsychological consequences that stem from sustaining a concussion. A growing base of structural and functional brain imaging studies indicate that adolescence is an extremely dynamic period of brain development [6,7], and a mTBI superimposed on a rapidly developing brain and body undergoing puberty reportedly leads to more severe and persistent symptoms in comparison to younger children and adults [8,9].

The current methods used in concussion detection, monitoring, and return-to-play clearance for adolescent athletes are suboptimal; these tools tend to lack either objectivity, utility, or feasibility for this population [10,11]. Functional brain imaging studies on concussion to date have begun to provide some clues regarding potential biomarkers of concussion. Resting-state fMRI studies have examined the disruptions to brain functional connectivity caused by concussion. This research has shown alterations in both dynamic (i.e., spending less time in a frontotemporal default mode/limbic brain state) and static measures of functional connectivity (i.e., altered interhemispheric connectivity, as well as hyperconnected frontal nodes and hypoconnected posterior nodes in the salience and fronto-parietal networks) relative to healthy control subjects [12,13].

While these findings are informative, the feasibility of performing multiple MRI tests on young athletes as an objective diagnosis/recovery assessment is not ideal and often not available in remote regions. Portable and feasible neuroimaging techniques are required that are sensitive to changes in the brain post impact. Electroencephalography (EEG) has the potential to serve as an objective diagnostic tool and has been extensively studied in recent years for this purpose.

EEG microstate analysis describes the discrete functional cortex-wide states that occur in the resting-state brain. During the resting state, the brain does not exist in one such state, but shifts dynamically between 4-7 different EEG “microstate topographies” that are usually stable for approximately 30-120 milliseconds (ms) before shifting to another state [14–16]. These microstates show high reliability [17]. This resting-state analysis method provides a lens for understanding altered dynamics of large scale brain networks, rather than isolated features of the EEG signal [18]. Importantly, EEG microstates have been shown to have strong associations with large-scale fMRI resting state brain networks (RSNs) [19]. In addition, EEG microstates capture subtle temporal dynamics about these functional brain areas and networks that cannot be captured with resting-state fMRI alone, given the limited temporal resolution of the BOLD signal. To our knowledge, EEG microstate analysis has been applied by only one group to the study of concussion [20]. However, this group investigated adults (mean age of 40 yrs) who had sustained a concussion several years prior (0.3 – 16 yrs) and there was no healthy control comparison group. In this chronic adult group with chronic neurophysiological impairement the duration of the four canonical microstates (A-D) were negatively correlated with a neuropsychological impairment index. Only four microstate classes were investigated and only average duration of each microstate was investigated. To our knowledge, this is the first study that has examined whether or not EEG microstate dynamics are altered in a adolecent sample in acute phase of mTBI injury.

In the present study, we investigate acute-phase changes in EEG microstate features in a cohort of adolescent males, with the primary aim to enhance our current understanding of neural dynamic changes associated with concussion. We hypothesized that mTBI will be associated with differences in microstate sequences as an indicator of acute changes in temporal dynamic interplay of large scale brain networks in comparison to healthy controls, and that these changes will be correlated with concussion symptoms. This pilot study is part of a larger project that aims to identify potential microstate markers of concussion to enhance the objectivity and reliability of concussion diagnoses and recovery assessments.

## Materials and methods

### Participants

Male athlete participants with normal or corrected to normal vision between the ages of 10 and 18 years were recruited for this study. Concussed participants received a diagnosis of concussion from a physician or team doctor. Any participants exhibiting focal neurologic deficits, pathology and/or those on prescription medications for neurological or psychiatric conditions were excluded from the study. All participants provided written assent and their parents gave written informed consent as per the guidelines of the Human Ethics Review Board of the University of British Columbia. The study was approved by the University of British Columbia Clinical Research Ethics Board (Approval number: H17-02973). The recruitment period of the study started on March 15, 2019 and is ongoing. The current ethics protocol was renewed and is valid until October 24, 2025. Eight of the concussed participants had completed either the Sport Concussion Assessment Tool 3 (SCAT3) or the Child SCAT3 (for children aged 12 and under), a standardized tool widely used for the evaluation of athletes suspected of having sustained a concussion [21].

### EEG data collection and preprocessing

Five minutes of eyes closed, resting-state EEG data were collected from individuals using a 64-channel HydroCel Geodesic Sensor Net (EGI, Eugene, OR). After obtaining assent and informed consent, participants were seated in an experimental room with controlled lighting levels and fitted with the EEG cap. They were instructed to minimize movement and remain seated with their eyes closed. Before initiating the data collection, the electrode-scalp resistance was ensured to be below 50 kΩ. The signals were referenced to the vertex (Cz) and recorded at a sampling rate of 250 Hz. The collected EEG data were imported into Python for preprocessing.

The initial step involved re-referencing the data from the current reference (Cz) to the average of all channels. This was followed by applying a 4th order Butterworth filter with zero phase shift for band-pass filtering, and a frequency range of 1 – 50 Hz was kept. This frequency range was chosen due to excessive noise at frequencies above 50 Hz, which interfered with subsequent preprocessing steps. Independent component analysis (ICA) was performed using the *infomax* algorithm and the MNE preprocessing package in Python [22]. To identify and remove noise-contaminated components, we employed the *ICLabel* algorithm, an automated labeling tool for independent components [22]. We retained only those components that the algorithm labeled as *“Brain”* (i.e. probability of the component presenting brain source was higher than artifactual source).

### Microstate analysis

For the microstate analysis, we utilized the *Pycrostates* package in Python [23]. Global Field Potential (GFP) peaks were extracted from individual data sets using the functions available in the *Pycrostates* package. This process includes calculating the standard deviation of all channel values at each time point, generating a time series of GFP values. Subsequently, peaks in this time series were identified using the *Scipy* integrated function, with the minimum distance between peaks set to the default value of 2 samples. We identified the minimum number of GFP peaks across all subjects. We then resampled the GFP series for each subject to match this minimum number. This step was performed to prevent the clustering results from being biased by variations in the number of peaks across individual data sets and to ensure uniformity. The GFP peaks were clustered using a modified k-means algorithm, aligning with established microstate literature.

The number of microstates used in previous studies typically ranges from 4 to 7 microstates; at present, there is no consensus on how to determine the optimal number of classes to use for EEG microstate analysis [19]. Although 4 canonical microstate templates (A to D) were initially prevalent in the EEG microstate literature, it has been argued that 7 distinct microstate templates (A to G) best capture the scope of spontaneous electrophysiological activity topographies that are observed in resting-state EEG studies [24]. We decided that 7 clusters optimally fit our cohort’s data, given that the 7-class solution maximized Global Explained Variance (GEV), and the alignment of the 7 identified microstates’ topographical shapes with extant literature [25].

We aligned the 7 microstates with the established microstate labels (A to G) in the literature, based on the topographical shapes of the states. We then assigned each time point of the original dataset to one of the 7 microstates, using a “winner takes all” strategy, a smoothing parameter of 6 samples (24 ms), and correlation between topographical maps as the similarity measure. This involved correlating each time point with each microstate and assigning the time point to the state with the highest correlation, taking smoothing parameter into account but not taking polarity into account. The smoothing was applied to reduce the influence of rapid fluctuations, potentially caused by artifacts, and to prevent false identification of microstate changes.

Three measures were extracted from each individual’s microstate sequence: average duration, occurrence rate, and time coverage. First, the average duration of each microstate was calculated by determining how long the microstate remained unchanged in each sequence. These individual durations were averaged for each subject, and group-level means, and standard deviations were extracted. The same method was applied to calculate the occurrence rate of each microstate (i.e., the number of occurrences per second) and the time coverage (i.e., the proportion of time each microstate occupied relative to the total recording time). Lastly, we also extracted the average transition rate per second between microstates for each individual, as the stability of staying in one state or frequently transitioning between states may be a characteristic of mTBI. Fig. 1 presents the pipeline. The original EEG time series is backfitted to these microstates using a “winner-takes-all” algorithm. Key metrics such as microstate duration, occurrence rate, and time coverage are then extracted for group comparison.

**Fig 1.**
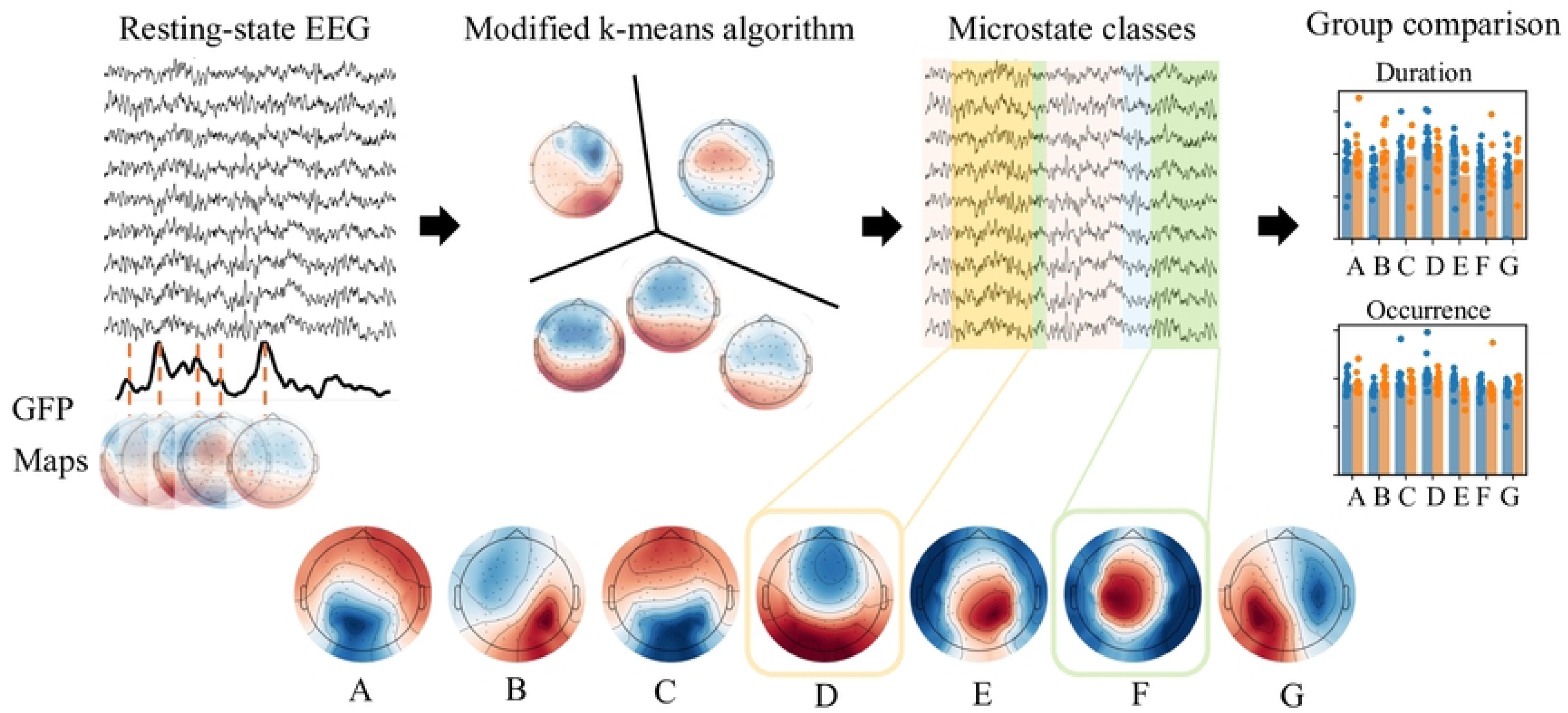
Study pipeline for microstate extraction from resting-state EEG to differentiate between groups. The Global Field Power (GFP) time series is computed, and microstate maps are identified by clustering maps from the local peaks of the GFP. Seven distinct microstates are identified. The maps are labeled based on their shape and the labeling method introduced in [25].

### Statistical analysis of microstate features

We used permutation statistics for all comparisons. For each feature, we conducted seven tests (one corresponding to each microstate). We applied a significance level of 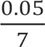 according to the Bonferroni method to adjust for multiple comparisons. During each test, we randomized the subjects from both groups and generated new groups of the same size as the original ones. We then conducted a t-test on these newly formed random groups. If the t-value from the random groups exceeded that of the original groups, we added the error rate by one. This procedure was repeated 10,000 times. The total error rate was then calculated and divided by the number of permutations. Features and microstates falling below the adjusted significance threshold are reported.

We conducted a multiple regression analysis to understand the relationship between symptom severity and quantity (as assessed by the SCAT) and microstate feature measures. Specifically, we used the mean duration and occurrence rate of each microstate as independent variables, while the reported quantity and severity of symptoms were used as the dependent variables. To account for multiple comparisons, the significance threshold was adjusted to 0.007. We did not include time coverage in this analysis, as it can be derived from the average duration and occurrence rate. If these two features show a significant relationship with symptoms, time coverage information is unnecessary.

## Results

### Demographics

34 male athletes were recruited: n = 14 with concussion and n = 20 with no history of concucssion. Data from two acutely concussed males were removed due to excessive noise remaining on EEG after preprocessing, leaving a final sample of 32 male adolescent athletes between the ages of 13 and 18 (Table 1). The total number of symptoms and symptom severity from the SCAT 3 were reported for 8 participants.

**Table 1.** Demographic and Clinical Characteristics of Study Participants.

| | Mean age (min:max) | SCAT total symptoms<br>(mean ± SD) | SCAT symptom<br>severity (mean ± SD) |
| --- | --- | --- | --- |
| Healthy Controls (N = 20) | 16.0 (14:18) | N/A | N/A |
| Acutely Concussed (N = 12) | 15.1 (13:18) | 9.1 ± 6.8 ($n = 8$) | 21.0 ± 20.4 ($n = 8$) |

### Microstate duration, occurrence rate and time coverage

Seven microstates with topographies similar to those identified in the meta-analysis by Koenig et al. (2024) were extracted from each subject (Fig. 1). The variance explained by each microstate series was calculated for individual datasets following the backfitting process (Fig. 2B). The total global explained variance (GEV) was 58 ± 12%. The analysis revealed several significant differences. First, microstate E was significant shorter in the concussed cohort compared to controls (*p* < 0.001) as shown by significantly lower duration, occurrence rate and time coverage of this microstate in the concussed group (*Cohen’s D* = 1.26, 1.08, 1.24 respectively). Additionally, microstate B exhibited a significantly higher mean duration, occurrence rate, and time coverage (*p* = 0.003, 0.003, < 0.001; *Cohen’s D* = 1.06, 1.07, 1.14 respectively) in the concussed group, i.e. there were fewer time points where the topographical map of the concussed data resembled microstate E, and more time points where it resembled microstate B (Fig.2B, C and D). We visualized these findings in two ways. In Fig. 2B, the average duration of microstates B and E is shown for each participant as individual data points, and by a bar plot comparing the group averages. In Fig. 3, we present the distribution of the duration for microstates B (Fig. 3 left) and E (Fig. 3 right). Specifically, each time one of these microstates occurred, its duration was recorded across data from all subjects. We then tested whether the distributions of these duration values significantly differed between the two cohorts using the Kolmogorov-Smirnov test and observed significant differences (p < 0.001).

**Fig. 2.**
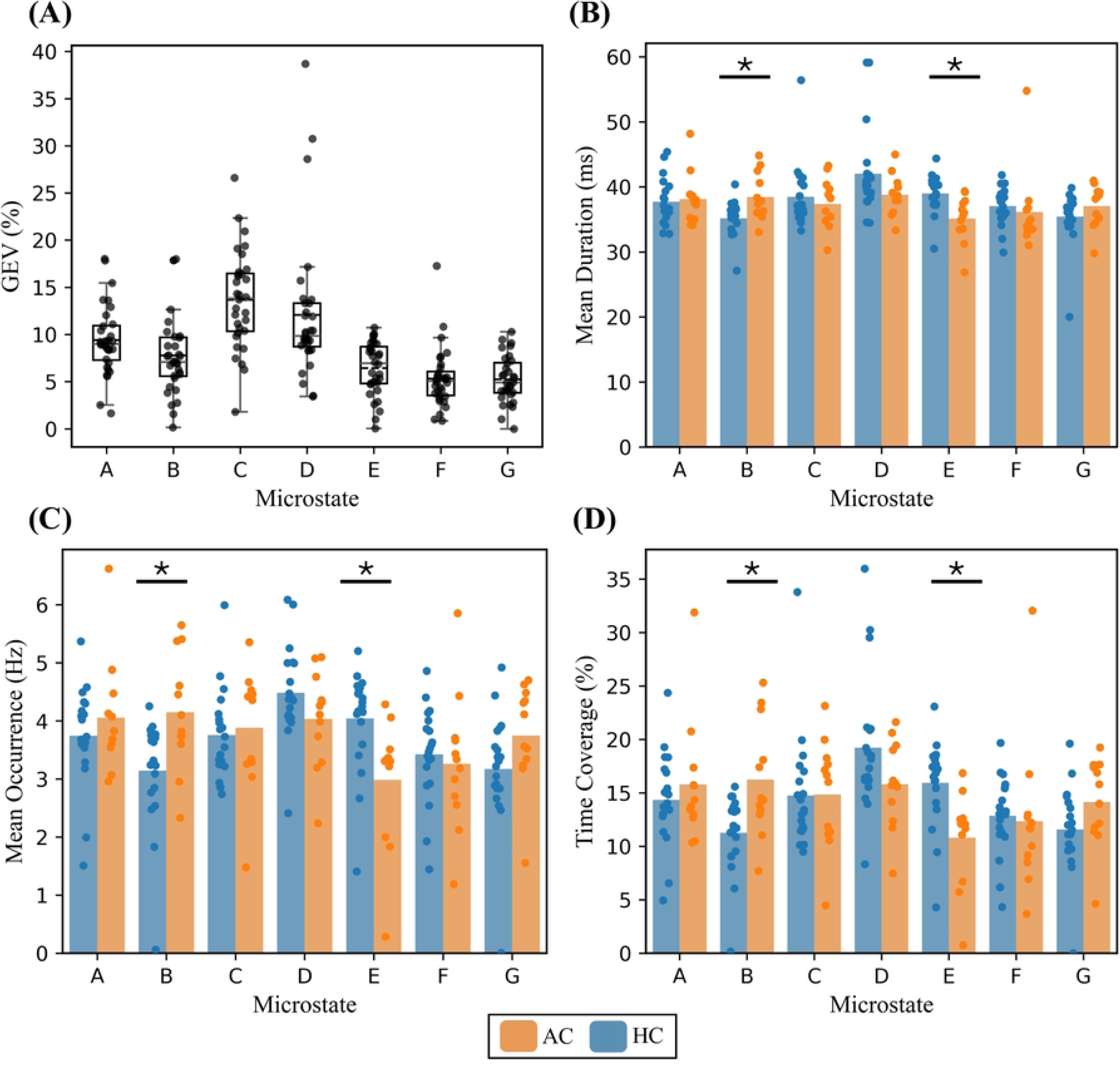
Microstate global variance, and comparative metrics in healthy vs. concussed participants. (A) Total global explained variance (GEV) of each microstate sequence, with each dot representing individual datasets. (B), (C) and (D) Bar plots representing the three key microstate measures: average duration, average occurrence rate, and time coverage, respectively. Healthy (blue, n = 20) and concussed (orange, n = 12) groups are compared, with dots representing individual data points. Significant differences in mean duration, occurrence rate, and time coverage of microstates B and E are observed. Overall, microstate E had a significantly lower duration, occurrence rate and time coverage in the concussed group compared to controls (*p* < 0.001). Additionally, microstate B exhibited a significantly higher mean duration, occurrence rate, and time coverage in the concussed group (*p* = 0.003). AC = Acute concussed; HC = Healthy Controls.

**Fig 3.**
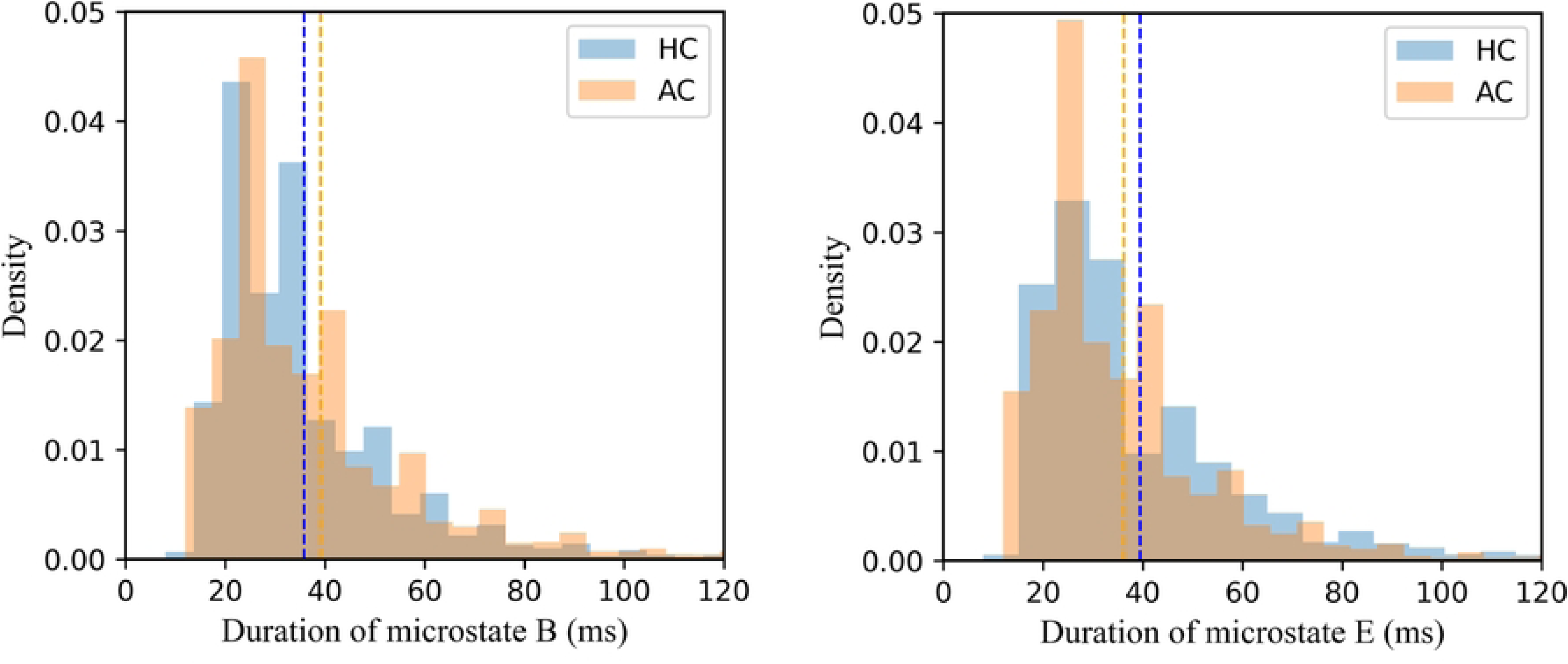
Distribution of microstate B and E durations across all occurrences. The distributions for microstate B and E durations are shown for both the HC (healthy control) and AC (acutely concussed) groups. Significant differences were observed between the duration distributions, with p < 0.001 (D = 0.01 for microstate B, D = 0.06 for microstate E).

### Microstate transition rate

Transitions were recorded for each second of the data. The total number of transitions per second was averaged over the entire data length for each individual. Interestingly, we observed higher variability in the number of transitions among the control group, though their average and median values were lower compared to the concussed group. However, this difference was not statistically significant (Fig. 4).

**Fig 4.**
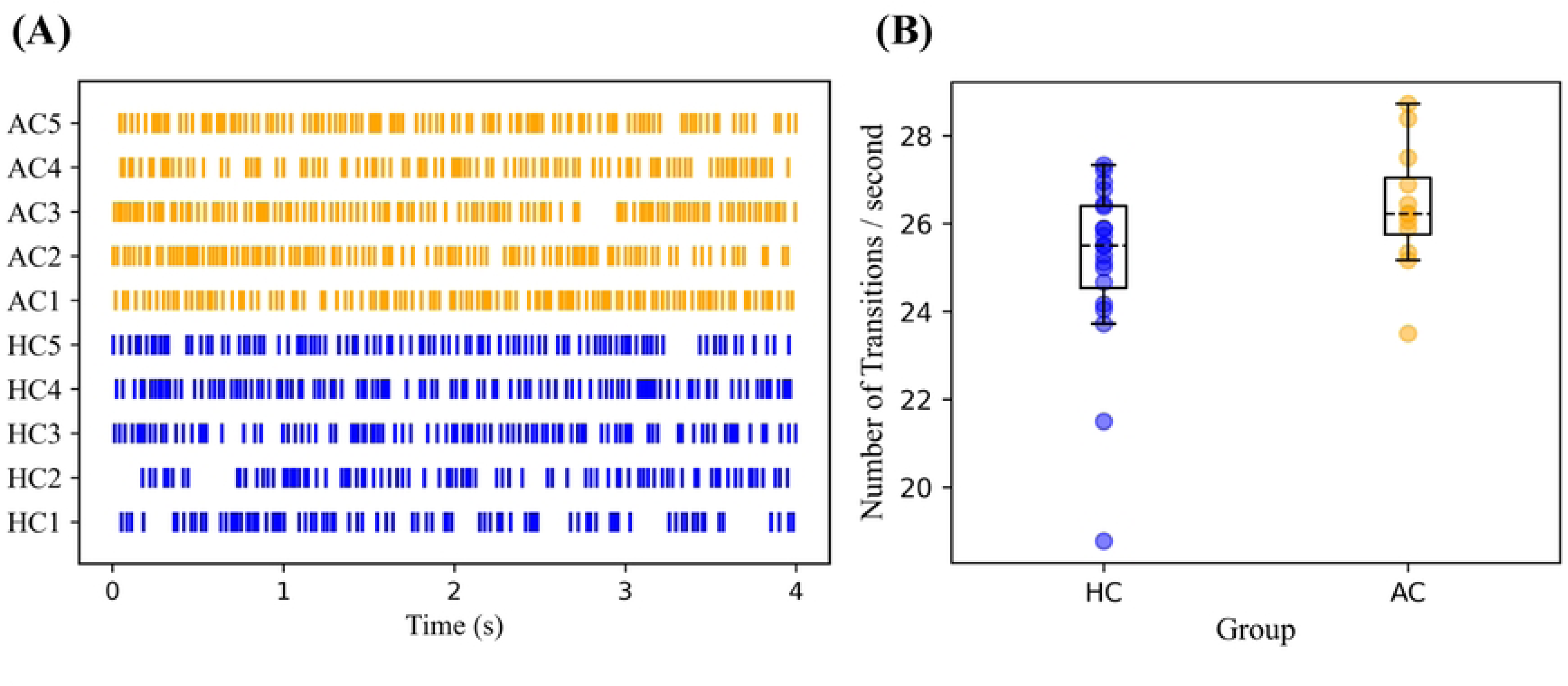
Rate of transitions in microstate sequences. (A) Shows a spike for each transition from one microstate to another within a 4-second time window for example subgroups. (B) Shows a box plot comparing the transition rates per second between the healthy control (HC) and acutely concussed (AC) groups, with individual data points representing the average number of transitions per second for each participant in the cohort. The difference between the two groups was not statistically significant.

**Fig 5.**
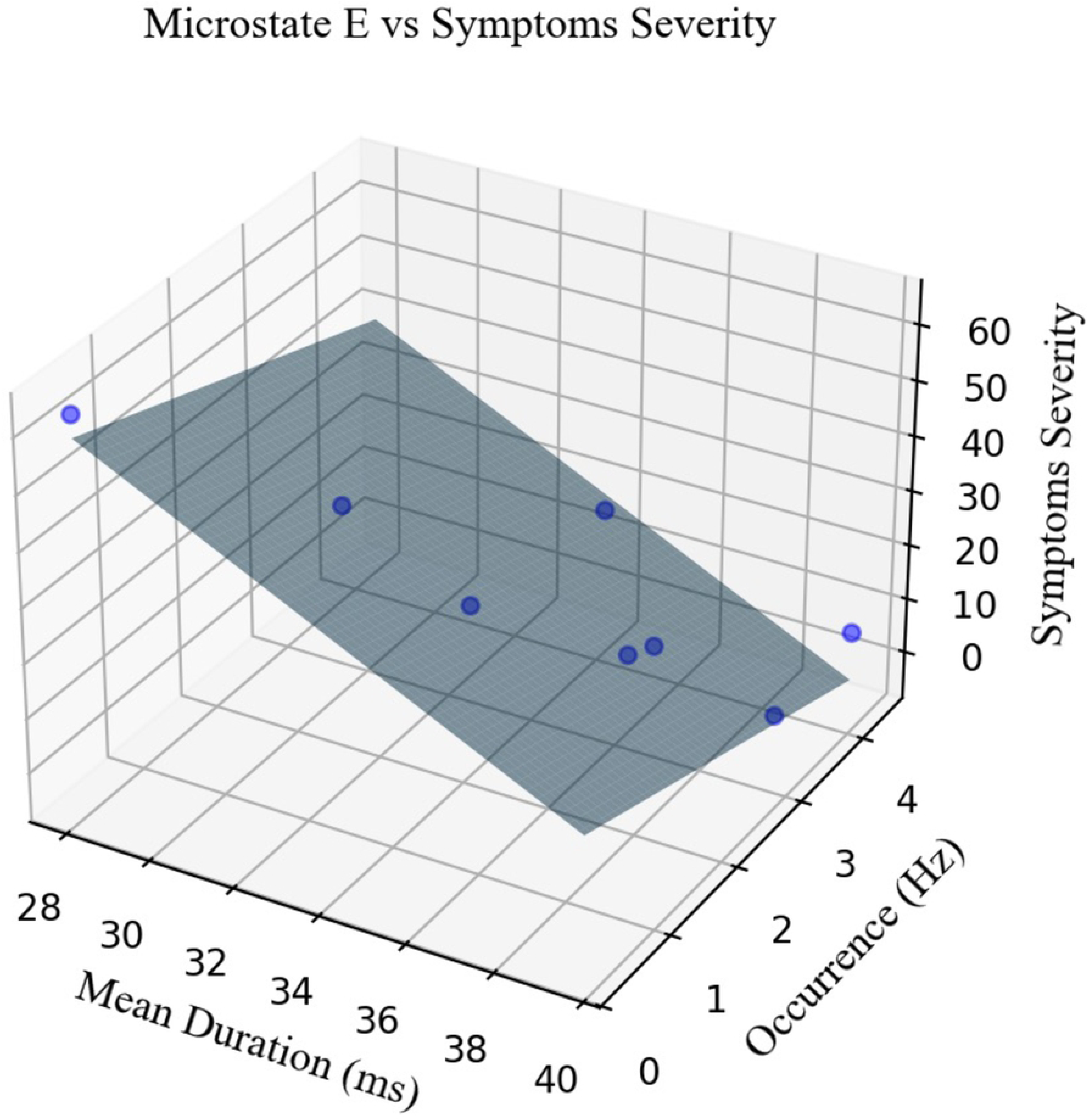
Multiple regression model. The average duration and occurrence rate of microstate E are used as independent variables, and symptom severity is used as the dependent variable. Data from eight concussed participants were included in this model due to the absence of SCAT information from other concussed individuals. The model demonstrates a statistically significant linear relationship between the duration and occurrence rate of microstate E and the reported symptom severity (*p = 0.006, F = 15.72*).

### Concussion symptoms related to microstate features

Among the eight concussed athletes who completed the SCAT, microstate E had a significant negative linear relationship with the SCAT symptom severity measure (*p = 0.006, F= 15.72*). This model indicates that increased symptom severity is associated with shorter duration and lower occurrence rates of microstate E.

## Discussion

In the present study, we applied EEG microstate analysis to study the changes associated with acute sport-related concussion in a small cohort of adolescent male athletes.

As hypothesized, we observed disturbed functional dynamics in the resting-state brain microstate of concussed adolescent participants relative to healthy control adolescent participants. Our findings show specific changes in microstates E and B: there was a significant decrease in the presence of microstate E and a significant increase in the presence of microstate B in our concussed sample across three microstate measures examined. Specifically, microstate B occured more frequently, lasted longer, and as a result covered a higher proportion of the total time; in contrast, this pattern was the opposite for microstate E. Interestingly, we also observed that the decreased occurrence rate and average duration of microstate E was associated with increased symptom severity within the concussed group. We did not find any significant differences in microstates transition rate between the groups.

EEG microstate analysis offers insights into temporal changes in the spatial organization of brain activity during rest. Specifically, this type of analysis relies on dynamic patterns of brain connectivity as opposed to a static measure of brain activity. Essentially, resting-state EEG scalp topographies tend to aggregate into a limited number of prototypical spatial distributions. These distributions are replicable both within and across individuals and have been extensively studied in the literature [17,25]. Typically, these topographical distributions are labeled A to G based on their maps. While labeling systems may vary across studies, we compared our results with those from studies that utilized similar topographical maps, though these studies report different labels. Our labeling system is based on the unifying work of Tarailis et al. [26] which facilitates cross-study comparisons.

Our findings indicate that the concussed group exhibits a reduced duration, occurrence rate and time coverage of microstate E. Microstate E is characterized by a topography with a centro-parietal maximum [26] and has been associated with activity in the default mode network (DMN) and the salience network (SN) [24,26]. Additionally, the scores of symptom severity showed a significant negative correlation with the duration and occurrence of this microstate. Given the critical role of the networks associated with microstate E in modulating both external and internal attention, we also observed a similar relationship in the concussed group where symptom severity which was linked to acute cognitive and somatic impairments were associated with characteristics of microstate E. Previous research has shown that sleep deprivation [27] and acute drug intoxication [28] are similarly associated with higher transition probability from microstate E to other microstates and reduced duration and coverage of microstate E respectively. Self-reported comfort at rest, on the other hand, has been associated with an increased duration of microstate E, which may be due to increased self-referential processing associated with the DMN [29]. These findings align well with what we observe in duration, occurrence and time coverage of microstate E. Although not statistically significant, we also observed a decrease in the duration, occurrence rate and time coverage of microstate D in the concussed group relative to the healthy control group.

Microstate D has been shown to primarily be associated with the frontoparietal executive control network (ECN) (e.g., activity in the right inferior parietal lobe and the right middle and superior frontal gyri) [24,30]. Given that the salience network is thought to modulate the dynamic interaction between the ECN and DMN [32], it is interesting that we observed a reduced presence of both microstates E and D in the concussed cohort, indicating potential impairment in key resting state networks related to attention regulation and cognitive control. Finally, a reduced duration of microstate E is consistent with the results of resting-state fMRI studies that have studied disruptions to dynamic functional connectivity in the brain as a result of concussion. Concussed pediatric participants have been observed to spend less time in a frontotemporal default mode/limbic brain state relative to their healthy control peers [13].

In contrast, microstate B characterized by a topography with a left frontal to right posterior configuration has been shown to be associated with the visual processing regions of the brain and particularly of activity in the left and right occipital cortices, including primary visual cortex [24,26]. It is possible that an altered balance of sensory processing activity in the brain (i.e., more activity in the visual imagery RSN), as a result of concussion, relates to acute cognitive impairments associated with concussion. Future research should explore, how concussion intervention alters the processing of various sensory inputs in the brain, and whether or not an imbalance in brain activity related to the processing of different sensory inputs (e.g., compensatory hyperactivity in one sensory modality) is a cause or effect of common concussion symptoms such as mental fatigue, attentional deficits, and memory impairment. It is also worth noting that in the aforementioned sleep deprivation study [27] and acute drug intoxication study [28], not only was there a decreased presence of microstate E, but also a higher transition probability from microstate E to microstate B. Perhaps concussion creates a state of impairment that prompts a similar neurobiological compensatory response to sleep deprivation and alcohol intoxication, where the brain is forced to prioritize bottom-up sensory processing (i.e., microstate B) over internally-oriented networks or higher-order top-down cognitive networks (i.e., microstates E and D) [28].

We also analyzed the number of transitions between microstates for each participant. This measure reflects the brain’s ability to switch between different functional states, which is crucial for maintaining efficient cognitive functioning. In a previous fMRI study, we observed reduced transitions between functional states in individuals with a concussion, suggesting cognitive rigidity [33]. However, we did not replicate this finding in the current study. A higher transition rate could potentially indicate neural instability or increased metabolic cost, which may also be linked to concussion pathology. Nevertheless, this observation did not reach statistical significance. To draw more definitive conclusions, future studies should explore this further with larger sample sizes and greater statistical power.

### Limitations and methodological considerations

Our study results have limited generalizability as we were only able to include a small cohort of male adolescent participants. Future studies will build upon the results of the present study by performing microstate analysis on a larger and more diverse sample and in a longitudinal design to address normal within-subject variabilities. An additional limitation is that only 8 of the 12 concussed subjects had completed the SCAT3, reducing the power of the correlation analysis performed between SCAT symptom quantity/severity scores and microstate features.

### Conclusion

In conclusion, using resting-state EEG in conjunction with microstate analysis, we observed significant disruptions in the dynamic interplay of large-scale brain networks in acutely concussed adolescent male athletes compared to their healthy control peers. EEG microstates provide a readily accessible and replicable method to study the effects of concussion on whole-brain network dynamics with high temporal resolution. This study aids in the effort to identify an objective biomarker for concussion, which is direly needed for the most accurate diagnosis and monitoring of this subtle and multifaceted injury.

## Data Availability

Data cannot be shared publicly because the data has been obtained from children. Data may be available for researchers who meet the criteria for access to confidential data by contacting the senior author (NVB -).

## Acknowledgements

We would like to express our gratitude to the participants of this study, whose cooperation and commitment made this research possible.

